# Platelet findings in 22q11.2 Deletion Syndrome correlate with disease manifestations but do not correlate with GP1b surface expression

**DOI:** 10.1101/2022.06.10.22276258

**Authors:** Ian M. Campbell, T. Blaine Crowley, Chintan Jobaliya, Alice Bailey, Daniel E. McGinn, Kimberly Gaiser, Anne Bassett, Raquel E. Gur, Bernice Morrow, Beverly S. Emanuel, Aime T. Franco, Deborah French, Elaine H. Zackai, Donna M. McDonald-McGinn, Michele P. Lambert

**Author notes:** **Address Correspondence to:** Michele P. Lambert, MD, MSTR, Division of Hematology, The Children’s Hospital of Philadelphia, 3516 Civic Center Blvd, Rm 316 G, Philadelphia, PA 19104.

## Abstract

Prior studies have demonstrated that patients with chromosome 22q11.2 deletion syndrome (22q11.2DS) have lower platelet counts (PC) compared to non-deleted populations. They also have an increased mean platelet volume. The mechanism for this has been postulated to be haploinsufficiency of the *GPIBB* gene. We examined platelet parameters, deletion size and factors known to influence counts, including status of thyroid hormone and CHD, in a population of 825 patients with 22q11.2DS. We also measured surface expression of GPIB-IX complex by flow cytometry. The major determinant of PC was deletion status of *GP1BB*, regardless of surface expression or other factors. Patients with nested distal chromosome 22q11.2 deletions (those with *GP1BB* present) had higher PCs than those with proximal deletions where *GP1BB* is deleted. Patients with 22q11.2DS also demonstrated an accelerated PC decrease with age, occurring in childhood. These data demonstrate that genes within the proximal deletion segment drive PC differences in 22q11.2DS and suggest that PC reference ranges may need to be adjusted for age and deletion size in 22q11.2DS populations. Bleeding did not correlate with either platelet count or GP1b expression. Further studies into drivers of expression of GP1b and associations with severe thrombocytopenia and immune thrombocytopenia are needed to inform clinical care.

## Introduction

The low copy repeat (LCR) mediated^1^ chromosome 22q11.2 deletion results in highly variable phenotypes.^2,3^ Although most (84%) patients with 22q11.2DS have the same size deletion caused by unequal crossing over between LCRs denoted LCR22A and LCR22D, the remaining patients have deletions nested within LCR22A and LCR22D.^2^ Most 22q11.2 deletions contain the *GP1BB* gene.^4^ Biallelic loss of function variants of *GP1BB* cause Bernard-Soulier Syndrome (BSS), a severe platelet disorder associated with macrothrombocytopenia. Individuals with 22q11.2DS with a second deleterious variant of *GP1BB* manifesting BSS have been reported.^5^

Multiple cytopenias have been reported in association with 22q11.2DS including: immune thrombocytopenia (ITP)^6^, autoimmune hemolytic anemia^7^ and Evans syndrome^8^ suggesting an increased risk of autoimmune cytopenias associated with the immune dysfunction present in some patients.^9^ However, in many cases there is mild, progressive thrombocytopenia not typical of ITP.

The prevailing hypothesis, that *GP1BB* haploinsufficiency drives 22q11.2DS associated thrombocytopenia, does not explain the variability in platelet count observed within this sub-population. We examined a large cohort of patients with 22q11.2DS to determine platelet characteristics towards helping guide management and better informing our understanding of platelet biology.

## Materials and Methods

The Institutional Review Board (IRB) at the Children’s Hospital of Philadelphia approved the longitudinal study of patients with a confirmed chromosome 22q11.2 deletion. Laboratory values and reference ranges were obtained from1,430 participants’ electronic health records, of whom 287 had whole genome sequencing. A subset of 78 patients were enrolled in an independently approved IRB protocol studying pediatric patient platelet biology. Of these, 34 had bleeding scores available. Participants/families provided separate consent for flow cytometric evaluation of platelet surface glycoprotein expression, evaluation of bleeding manifestations, and history of surgical bleeding. Please see supplementary materials for complete methods.

## Results and Discussion

### Platelet Indices

Platelet counts in patients with 22q11.2DS are not static, but decrease over time (**Figure 1A**). Similar to data in a non-deleted population, ^10^ we demonstrate a decline in platelet count with age that is accelerated to childhood and a gender difference showing instead that males have lower platelet counts (**Figure 1A**) (p=0.018) without significant gender differences in MPV (**Figure 1B**).

**Figure 1:**
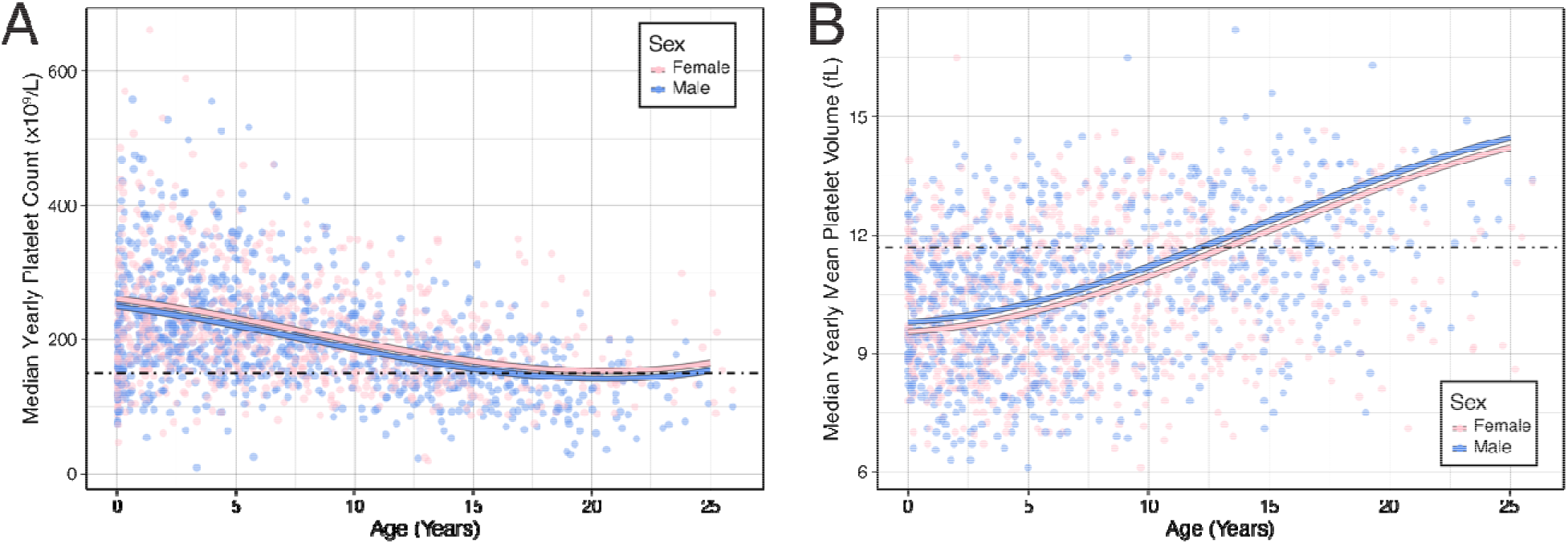
Platelet count and MPV versus age in 22q11.2DS. **A**. Median platelet count for each patient during that year of life for female (pink) and male (cyan) individuals. Dashed line: lower limit of normal. The colored lines: mean of the linear mixed-effects regression model for that sex. **B**. As (A) but for MPV. Dashed line: upper limit of normal.

Immune thrombocytopenia is a reported association with 22q11.2DS.^3,9^ Although we saw IPF increase during the second decade of life in individuals with 22q11.2DS (**Supplementary Figure – SF1**), we did not observe any correlations between platelet count and immunoglobulin levels or lymphocyte counts (data not shown).

### Influence of Thyroid Function and CHD

Thyroid dysfunction, associated with abnormalities in platelet counts,^11^ is a common medical issue in 22q11.2DS^2^. We examined the interaction between thyroid hormone levels and platelet count and found that clinical or subclinical hypothyroidism may partially explain low platelet counts in our participants (**SF2**), potentially through a direct effect of T4 on megakaryocyte proliferation (**SF2-E**). There was a correlation between CHD severity and platelet count (**SF-2F**).

### Haploinsufficiency of GP1BB

A hypothesis frequently noted in the literature for alterations in platelet indices in patients with 22q11.2DS is haploinsufficiency of *GP1BB*. Some data conflicts with prior small studies, such as by Brenner et al.^14^ suggesting that GP1b expression did not explain changes in platelet counts, and others suggesting haploinsufficiency of *GPIBB* does not explain platelet count changes^15^. As noted above, some patients with 22q11.2DS have deletions nested within *LCR22A-LCR22D* (**SF3**). Those deletions excluding the *LCR22A-LCR22B* segment are expected to have two copies of *GP1BB*. Therefore, we assessed whether deletion size (*GP1BB* haploinsufficiency) influenced platelet indices. We found patients with *LCR22A-LCR22B* deletions had average decreased platelet counts of 80×10^9^/L compared to individuals with 22q11.2 deletions excluding *LCR22A-LCR22B* (p < 1×10^−9^, **Figure 2A)** as well as 1.3 fL larger MPV on average (p = 2.8 × 10^−4^, **Figure 2B**).

**Figure 2:**
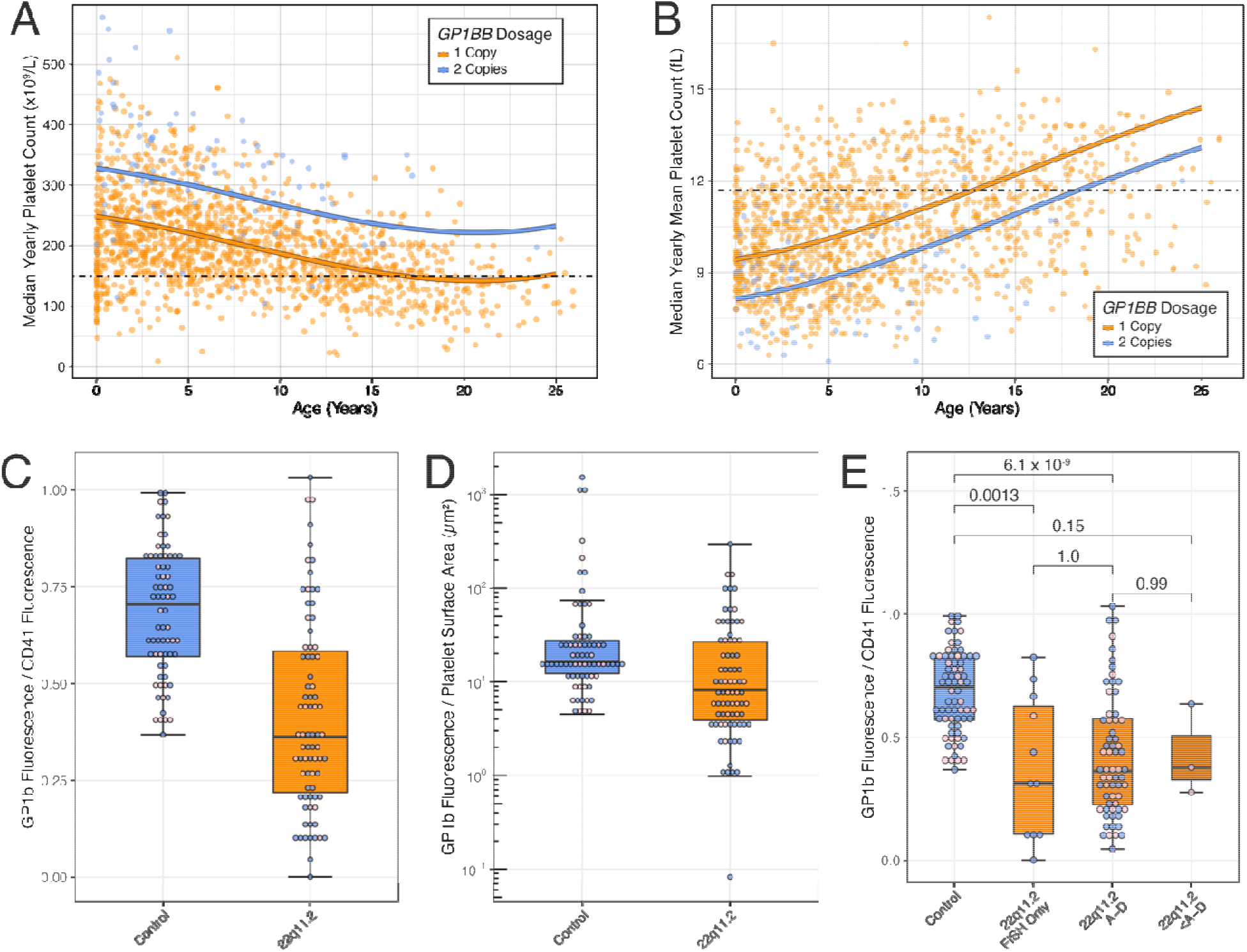
Deletion of *LCR22A-LCR22B* status associated with platelet parameters and GP1b expression. **A**. Deletion of *GP1BB* versus platelet count and age. The orange line: mean value of the linear mixed-effects regression model for deletion including *LCR22A-LCR22B* encompassing *GP1BB; b*lue line: deletion excluding the *LCR22A-LCR22B*. Points indicate median platelet count individuals for a year. Individuals tested by FISH are expected to have the A-B region deleted, but the status of other inter-LCR regions is unknown. B. Same as (A) for MPV. **C**. Ratio of expression of GP1b:CD41 in control subject (blue) versus subjects with 22q11.2DS (orange) whiskers represent minimum and maximum (excluding outliers); middle line represents median; boxes represent interquartile range and grey dots represent individual values including outliers. **D**. As in (C) but for ratio of GP1b:platelet surface area as estimated by side scatter plot. **E**. As in (C) and separated by deletion size. P-values as Dunn post hoc analysis after a significant Kruskal Wallis test (p = 3.7 × 10^−9^) corrected for multiple testing with Benjamini and Hochberg method.

Our data, in a large cohort of patients with 22q11.2DS suggests that inclusion of *GPIBB* within the deleted region accounts for much of the variability in platelet counts and GP1b expression.

### Expression of Glycoprotein (GP) 1b

Surface expression of GP1b may be mediating low platelet count and high MPV. We found individuals with 22q11.2DS had significantly lower GP1b expression, as a ratio of expression compared to CD41 and as a function of estimated platelet surface area (**Figure 2C and Figure 2D**, p ≤ 0.8 × 10^−5^). As expected, the median expression of GP1b was almost exactly half that of controls (ratio 0.504). Deletion size did not contribute additionally to GP1b:CD41 ratio (**Figure 2E**), but likely because no patient with flow cytometry data had a deletion excluding *GP1BB* (see supplementary material for further details).

Although, GP1b expression is correlated with platelet count and inversely correlated with MPV across both cohorts (p = 0.031, p = 0.0012, respectively, **SF4**), expression level does not explain more variance than deletion status alone (p = 0.21, p = 0.45, respectively). Although 22q11.2 deletions encompassing *GP1BB* appear to cause approximately half normal levels of expression of GP1b, variation in expression of GP1b beyond that mediated by the deletion does not affect platelet indices.

### Comparison of bleeding with platelet count and expression of GP1b

Because either platelet count or surface expression of GP1b might influence risk of clinically significant bleeding, we assessed bleeding outcomes with a previously validated scoring system (see supplementary materials).^13^ However, we were unable to identify any significant correlations between bleeding score and platelet count or GP1b expression (p = 0.16 and 0.14, respectively**SF5**).

### Genetic Variation of GP1BB

Finally, to assess the impact of possible genetic variation of the remaining *GP1BB* gene, we utilized whole genome sequencing and CBC data available for 215 of 287 participants. No coding sequence variants of *GP1BB* were identified. None of the 415 identified variants within 10kb of the coding sequences and one intron of *GP1BB* were significantly associated with platelet count or MPV (addressing regulatory element variation). We found no statistically significant associations with common or rare variants in our cohort (see supplemental data).

## Conclusions

Our study demonstrates that platelet count in individuals with 22q11.2DS changes over time and that deletion of the *LCR22A-LCR22B* region harboring the *GP1BB* gene is clearly associated with decreased platelet count and increased mean platelet volume. Additionally, we identified an approximate 50% decrease in expression of GP1b in patients with 22q11.2DS. This decreased expression is associated with a modest decrease in platelet count and increase in MPV. Both thyroid hormone levels and CHD were correlated with platelet count independently but not with platelet indices. These associations were independent of patient age suggesting they also contribute to platelet count variability. Neither platelet count nor GP1b expression was clearly correlated with bleeding. Our data provide some potential additional mechanisms to account for variability in platelet counts, but further studies are needed to better understand the relationship between platelet count and bleeding risk as well as to delineate other determinants of platelet count to better counsel patients and families.

## Data Availability

Cardiac phenotype and sequencing data are publicly available in dbGaP: Data dictionaries and variable summaries are available on the dbGaP FTP site:   (ftp://ftp.ncbi.nlm.nih.gov/dbgap/studies/phs001339/phs001339.v1.p1), while the public summary-level phenotype data may be browsed at: (https://www.ncbi.nlm.nih.gov/projects/gap/cgi-bin/study.cgi?study_id=phs001339.v1.p1). Whole genome sequencing data is available through the International Brain and Behavior Consortium (http://www.22q11-ibbc.org). De-identified laboratory data is available upon request from D.M.M.-M.

ftp://ftp.ncbi.nlm.nih.gov/dbgap/studies/phs001339/phs001339.v1.p1

https://www.ncbi.nlm.nih.gov/projects/gap/cgi-bin/study.cgi?study_id=phs001339.v1.p1)

## Data availability

Cardiac phenotype and sequencing data are publicly available in dbGaP: Data dictionaries and variable summaries are available on the dbGaP FTP site: (ftp://ftp.ncbi.nlm.nih.gov/dbgap/studies/phs001339/phs001339.v1.p1), while the public summary-level phenotype data may be browsed at: (https://www.ncbi.nlm.nih.gov/projects/gap/cgi-bin/study.cgi?study_id=phs001339.v1.p1).

Whole genome sequencing data is available through the International Brain and Behavior Consortium (http://www.22q11-ibbc.org). De-identified laboratory data is available upon request from D.M.M.-M.

## Acknowledgements

The investigators gratefully acknowledge the participation of patients and families with 22q11.2DS. In addition, we thank Dr. Kathleen Sullivan and Dr. Vaneeta Bamba for their comments and suggestions with the analysis of the immunologic and endocrinologic data. This study was funded by the NIH: UO1MH191739, U01MH087636, PO1HD070454.

## Author Information

Conceptualization: I.M.C., T.B.C., A.B., R.E.G., B.M., B.S.E, A.F., E.Z., D.M.M.-M., M.P.L.; Data curation: I.M.C., T.B.C., D.E.M., K.G., A.B. D.M.M.-M., M.P.L.; Formal Analysis: I.M.C., T.B.C., C.J., D.F., D.M.M.-M., M.P.L.; Funding acquisition: A.B., B.S.E, D.M.M.-M., M.P.L.; Investigation: I.M.C., C.J., D.F., M.P.L.; Resources: A.B., E.Z., D.M.M.-M., M.P.L.; Visualization: I.M.C, C.J., D.F., M.P.L.; Writing – original draft: I.M.C., D.M.M.-M., M.P.L.; Writing – review & editing: I.M.C., T.B.C., D.M.M.-M., M.P.L.

## Ethics Declaration

The Institutional Review Board (IRB) at the Children’s Hospital of Philadelphia approved the longitudinal cohort study of patients with 22q11.2DS. Informed consent was obtained from the patient or their guardian for publication of data. Individual-level data was deidentified.

## References

1. Edelmann L, Pandita RK, Morrow BE. Low-copy repeats mediate the common 3-Mb deletion in patients with velo-cardio-facial syndrome. The American Journal of Human Genetics. 1999;64(4):1076–1086.

2. Campbell IM, Sheppard SE, Crowley TB, et al. What is new with 22q? An update from the 22q and You Center at the Children’s Hospital of Philadelphia. Am J Med Genet A. 2018;176(10):2058–2069.

3. McDonald-McGinn DM, Sullivan KE, Marino B, et al. 22q11.2 deletion syndrome. Nature Reviews Disease Primers. 2015;1:15071.

4. Kahn ML, Diacovo TG, Bainton DF, Lanza F, Trejo J, Coughlin SR. Glycoprotein V-Deficient Platelets Have Undiminished Thrombin Responsiveness and Do Not Exhibit a Bernard-Soulier Phenotype. Blood. 1999;94(12):4112–4121.

5. Budarf ML, Konkle BA, Ludlow LB, et al. Identification of a patient with Bernard-Soulier syndrome and a deletion in the DiGeorge/velo-cardio-facial chromosomal region in 22q11.2. Hum Mol Genet. 1995;4(4):763–766.

6. Lawrence S, McDonald-McGinn DM, Zackai E, Sullivan KE. Thrombocytopenia in patients with chromosome 22q11.2 deletion syndrome. The Journal of Pediatrics. 2003;143(2):277–278.

7. Cancrini C, Puliafito P, Digilio MC, et al. Clinical Features and Follow-Up in Patients with 22q11.2 Deletion Syndrome. The Journal of Pediatrics. 2014;164(6):1475-1480.e2.

8. Kratz CP, Niehues T, Lyding S, et al. Evans Syndrome in a Patient with Chromosome 22q11.2 Deletion Syndrome: A Case Report. Pediatric Hematology and Oncology. 2003;20(2):167–172.

9. Crowley TB, Campbell IM, Liebling EJ, et al. Distinct immune trajectories in patients with chromosome 22q11.2 deletion syndrome and immune-mediated diseases. Journal of Allergy and Clinical Immunology. 2022;149(1):445–450. doi:10.1016/j.jaci.2021.06.007

10. Bonaccio M, Di Castelnuovo A, Costanzo S, et al. Age- and sex-based ranges of platelet count and cause-specific mortality risk in an adult general population: prospective findings from the Moli-sani study. Platelets. 2018;29(3):312–315.

11. Naqvi N, Davidson SJ, Wong D, et al. Predicting 22q11.2 deletion syndrome: a novel method using the routine full blood count. Int J Cardiol. 2011;150(1):50–53.

12. Billett J, Cowie MR, Gatzoulis MA, et al. Comorbidity, healthcare utilisation and process of care measures in patients with congenital heart disease in the UK: cross-sectional, population-based study with case-control analysis. Heart. 2008;94(9):1194–1199.

13. Buchanan GR, Adix L. Grading of hemorrhage in children with idiopathic thrombocytopenic purpura. J Pediatr. 2002;141(5):683–688.

14. Brenner MK, Clarke S, Mahnke DK, et al. Effect of 22q11.2 deletion on bleeding and transfusion utilization in children with congenital heart disease undergoing cardiac surgery. Pediatr Res. 2016;79(2):318–324. doi:10.1038/pr.2015.216

15. Zwifelhofer NMJ, Bercovitz RS, Weik LA, et al. Hemizygosity for the gene encoding glycoprotein Ibβ is not responsible for macrothrombocytopenia and bleeding in patients with 22q11 deletion syndrome. Journal of Thrombosis and Haemostasis. 2019;17(2):295–305.

