## Supplemental Materials for "Platelet findings in 22q11.2 Deletion Syndrome correlate with disease manifestations but do not correlate with GP1b surface expression"

Supplementary Materials:

**Methods and Materials:**

*Sequencing*

Sequencing data and phenotype correlations for many patients within the cohort have been previously reported.^1^ Patients were sequenced using MLPA (Multiplex ligation-dependent probe amplification), SNP microarray, CGH (comparative genomic hybridization), or FISH (fluorescent *in situ* hybridization) as previously described.^2^ WGS with a median depth of 39-fold was performed on 287 subjects as part of the International 22q11.2 Brain and Behavior Consortium (which sequenced 1,595 subjects from 22 international clinical sites).^3^ In brief, samples were sequenced with the Illumina HiSeq X Ten for the first 100 samples and the Illumina HiSeq 2500 platform for all other samples at Hudson Alpha. Sequence reads were mapped to genome build hg38 (December 2013; GRCh38/hg38) with PEMapper (90% stringency for 2 × 100 bp reads and 95% stringency for 2 × 150 bp reads;).^4^

*Flow cytometry*

Study participant and control samples were generally matched by sex and age (<2 years, 2-6 years, 6-12 years, 12-18 years and >18 years). Comparison of platelet parameters with age was done using the validation cohort from the general hematology laboratory. Study participant samples were collected in a 2.7 ml sodium citrate tube after informed consent and at the same time as a clinical sample. Forty microliters of whole blood were resuspended in 800 mcl Dulbeccos phosphate buffered saline (DPBS) with 0.2% bovine serum albuin (BSA); then 20 mcl of the diluted was incubated in the dark with the following antibodies for 15 minutes: FITC-anti-CD42a, PE-anti-CD42b, PE-Cy7-anti-CD61 and ECD-anti-CD41 (all BD Biosciences, San Jose, CA). Staining for CD42a and CD61 were used to confirm platelet gates and ensure adequate staining with antibodies of interest: CD42b (GP1b) and CD41. Samples were then fixed with Cytofix buffer (1:10 ml; BD Biosciences) for 10 minutes and washed with DPBS with BSA and resuspended in 1 ml for flow cytometric analysis. Samples were processed and fixed within 2 hours of collection and analyzed within 24 hours of collection on an LSR II flow cytometer (BD Biosciences). Flow cytometric analysis was performed primarily on patients with the classical *LCR22A-LCR22D* deletion and only small numbers of patients with flow cytometric data had 22q11.2 deletion diagnosed by only by FISH (fluorescence in situ hybridization), which tested for the presence of the *TPL1* gene within the *LCR22A-LCR22B* region but does not inform the size of the deletion and 3 patients had smaller deletions all of which *did* span the *LCR22A-LCR22B* region at a minimum. There were no patients analyzed by flow cytometry who had deletions that did not encompass the *LCR22A-LCR22B* region.

*Platelet Indices Analysis*

Routine complete blood count data including platelet count and mean platelet volume (MPV) was analyzed clinically in the clinical hematology laboratory at The Children’s Hospital of Philadelphia on a Sysmex 6000N analyzer using standard parameters (Sysmex, Inc). Immature platelet fraction was collected from fluorescent platelet measurements in patients on whom platelet profile was clinically requested and compared to internally validated reference cohort data. For longitudinal analysis, we evaluated participants with at least 3 complete blood counts (CBCs) during the study period (n = 456), limiting to the median value during each chronological year for individuals with multiple CBCs. In contrast to CBCs, IPF measurement is not standard clinical practice in our center, and therefore considerably fewer participants had IPF measured at any point (n = 50) and there is likely bias in which patients were selected for IPF measurement.

*iPSC-derived Megakaryocyte Differentiation*

To evaluate the direct effect of thyroid hormone on megakaryopoiesis, human induced pleuripotent stem cells (iPSC) were differentiated to megakaryocytes with and without LT4 (thyroxine). An established control human iPSC line, designated CHOPWT10,^5^ was differentiated to hematopoietic progenitor cells (HPCs) using a previously published protocol.^6^ Cryopreserved HPCs were thawed, counted, and plated at 3 x 10^4^ cells/well of a 24-well Ultra-low attachment tissue culture plate in StemSpan SFEM II (Stem Cell Technologies) medium at 37°C in normoxic conditions. Cells were resuspended in medium containing the cytokines, TPO (50 ng/ml) and SCF (25 ng/ml), for expansion to megakaryocytes and preparations of human thyroxine (T4) hormone at 25 nM, 50 nM, 100 nM, and 200 nM respectively. All conditions were tested in triplicate. Cells were fed by adding medium to the wells on days 2 and 4 of incubation. On day 5 of incubation, cells were harvested, counted, and flow cytometry was performed for the megakaryocyte markers CD41 and CD42b (GP1b) as above.

*Bleeding evaluation*

Patients enrolled in the platelet biology study (n= 78) had a bleeding score assessed by review of medical records (modified Buchannan and Adix score as previously described)^7^. These scores were correlated with platelet count and platelet surface marker expression. There were 2 patients with known Immune Thrombocytopenia (ITP) and one patient with concomitant vonWillebrand Disease confirmed on hematology evaluation.

*Statistical Analysis*

Data analysis and statistical testing were performed using the R Statistical Programing Language. To assess for association between platelet indices and various patient parameters, statistical models were developed. For data modeled against time, such as platelet count and MPV, a third order polynomial fixed effect of age was employed. For data with repeated measures, linear mixed-effects models were created using both a subject and subject-age interaction random effect (lme4 package). This limited over-contribution of a single subject being measured multiple times. For models without a time component or repeated measures, a generalized linear model was used. For analysis of bleeding score, both Pearson correlation and ordinal logistic regression (ordinal package) approaches were attempted. To assess difference of continuous variables between two groups, the Wilcox signed rank test was employed. To assess multiple groups, the Kruskal Wallis omnibus test was employed with a Dunn test post hoc analysis corrected in the manner of Benjamini and Hochberg (FSA package). For genotype comparisons, Bonferoni correction was used to control for multiple testing.

**References**

1.	Mlynarski EE, Xie M, Taylor D, et al. Rare copy number variants and congenital heart defects in the 22q11.2 deletion syndrome. *Hum Genet*. 2016;135(3):273-285.

2.	Cohen JL, Crowley TB, McGinn DE, et al. 22q and two: 22q11.2 deletion syndrome and coexisting conditions. *Am J Med Genet A*. 2018;176(10):2203-2214.

3.	Gur RE, Bassett AS, McDonald-McGinn DM, et al. A neurogenetic model for the study of schizophrenia spectrum disorders: the International 22q11.2 Deletion Syndrome Brain Behavior Consortium. *Mol Psychiatry*. 2017;22(12):1664-1672.

4.	Johnston HR, Chopra P, Wingo TS, et al. Reply to Pluss et al.: The strength of PEMapper/PECaller lies in unbiased calling using large sample sizes. *Proc Natl Acad Sci U S A*. 2017;114(40):E8323.

5.	Maguire JA, Gagne A, Mills JA, Gadue P, French DL. Generation of human control iPS cell line CHOPWT9 from healthy adult peripheral blood mononuclear cells. *Stem Cell Res*. 2016;16(1):14-16.

6.	Mills JA, Paluru P, Weiss MJ, Gadue P, French DL. Hematopoietic differentiation of pluripotent stem cells in culture. *Methods Mol Biol*. 2014;1185:181-194.

7.	Buchanan GR, Adix L. Grading of hemorrhage in children with idiopathic thrombocytopenic purpura. *J Pediatr*. 2002;141(5):683-688.

**Supplemental Data:**

*Supplemental genetic analysis data*

While hemizygosity for the common variant, rs35098261, was initially associated with both decreased platelet count and increased MPV this was not significant after correcting for multiple testing (n = 14). We found no associated common variation at the *TUBB1* and *ACTN1* loci (independently associated with platelet values).

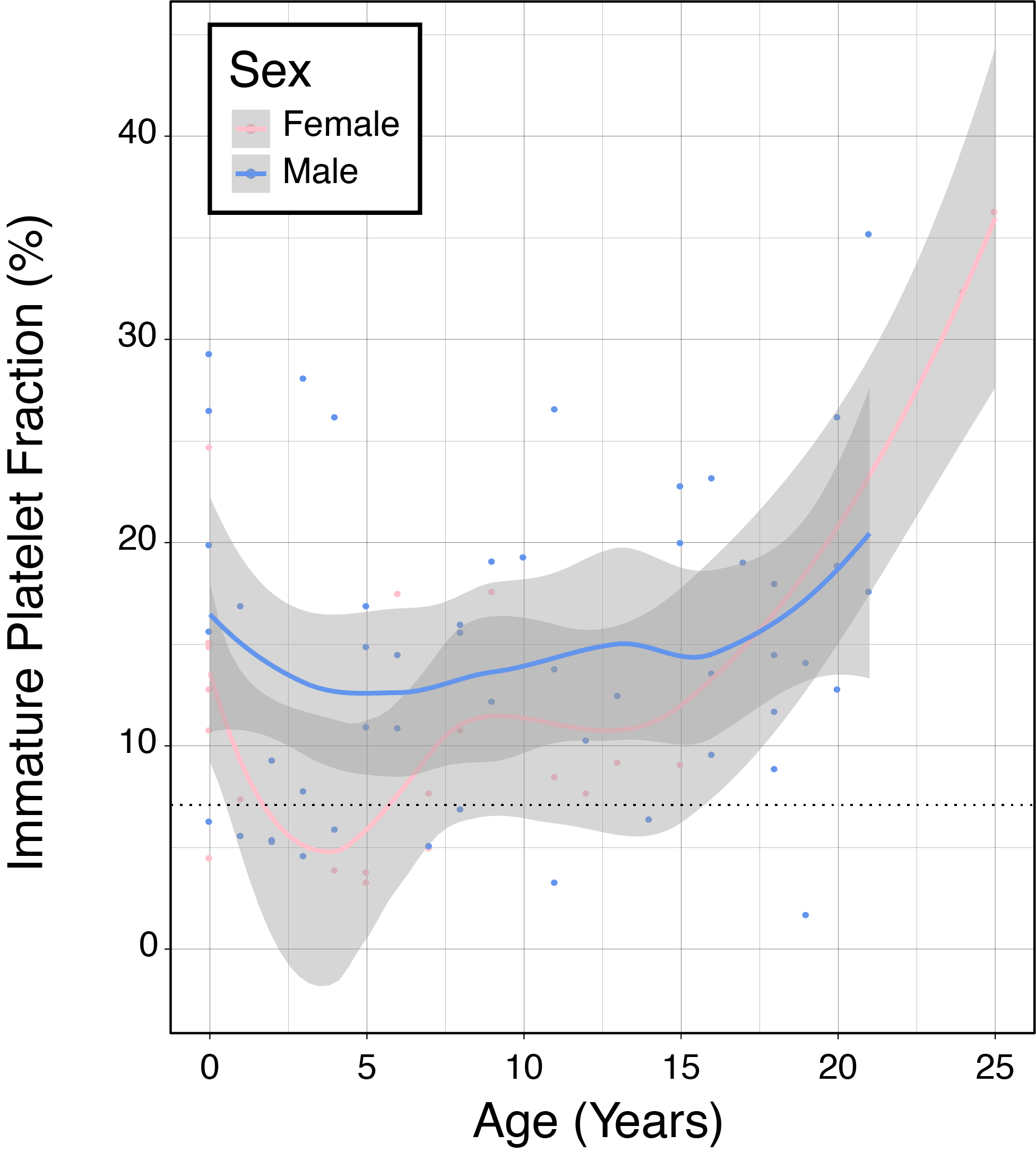

**Supplementary Figure 1:** Immature platelet fraction over time among individuals with 22q11.2DS. Pink line (LOESS fit) and dots (individual values) are female individuals while blue represents males. Grey areas represent 95% confidence interval of the mean.

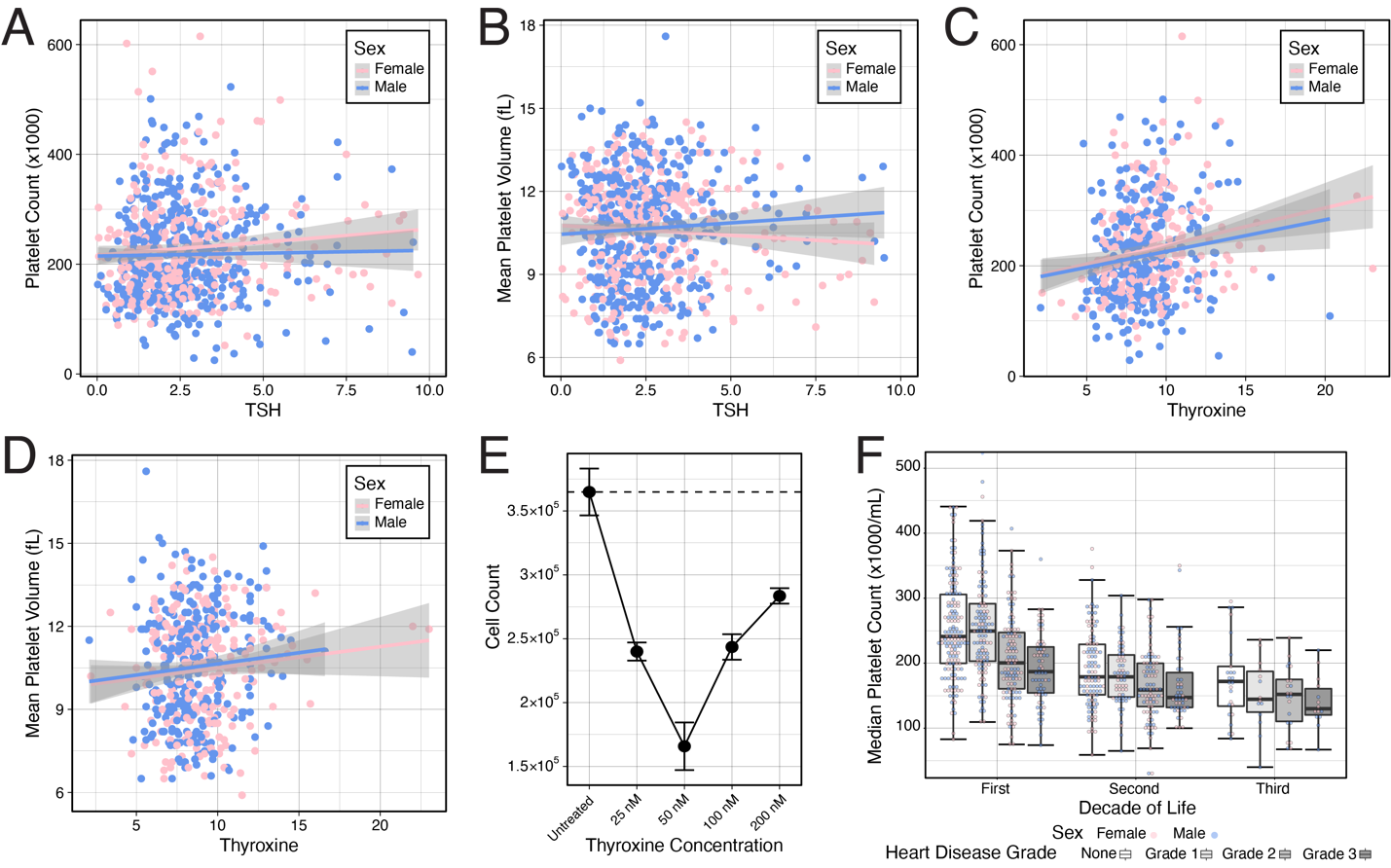

**Supplementary Figure 2:** Platelet count and MPV related to thyroid hormone and congenital heart disease severity. **A**. Platelet count vs simultaneous thyroid-stimulating hormone (TSH) measurement for female (pink) and male (cyan) individuals. The colored lines indicate a generalized linear model fit or that sex with 95% confidence interval (gray channels). **B**. As (A) but for MPV and TSH. **C.** As (A) but for PC and thyroxine (T4). **D.** As (A) but for MPV and T4. **E.** iPSC derived megakaryocytes on day 5 of differentiation versus T4 (x-axis). Broken line indicates control cell numbers. n=3, p<0.001. **F.** Congenital heart disease severity (varying shades of grey boxes) and by decade of life. Whiskers represent minimum and maximum (excluding outliers); middle line represents median; boxes represent interquartile range. Dots represent individual values (including outliers) male (blue) and female (pink) individuals.

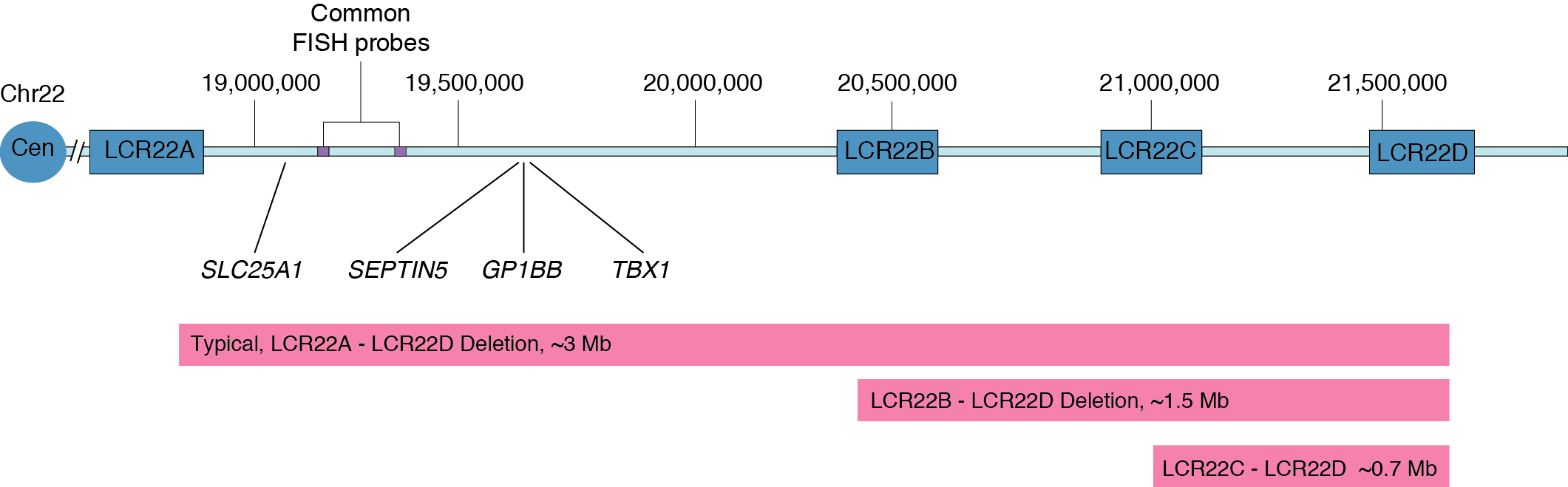
**Supplementary Figure 3**: Schematic representation of the organization of low copy repeats and notable genes in the 22q11.2 region.

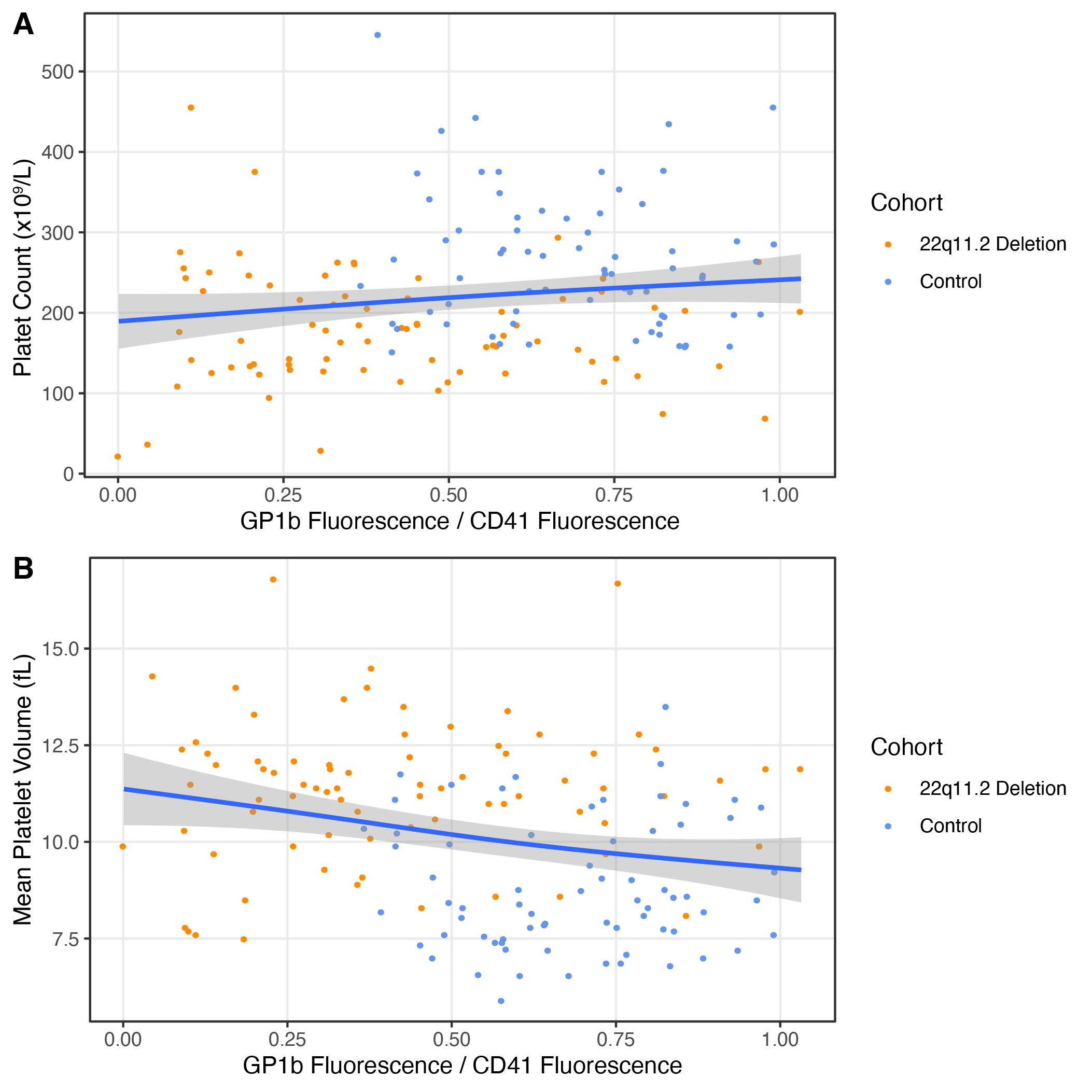

**Supplementary Figure 4**: Platelet parameters versus GP1b expression. (A) Ratio of

GP1b/CD41 expression versus platelet count for patients with 22q11.2DS (orange dots) versus control (blue dots) individuals. Blue line represents a linear model fit and grey area is 95% confidence interval. (B) as in (A) but for Mean Platelet Volume.

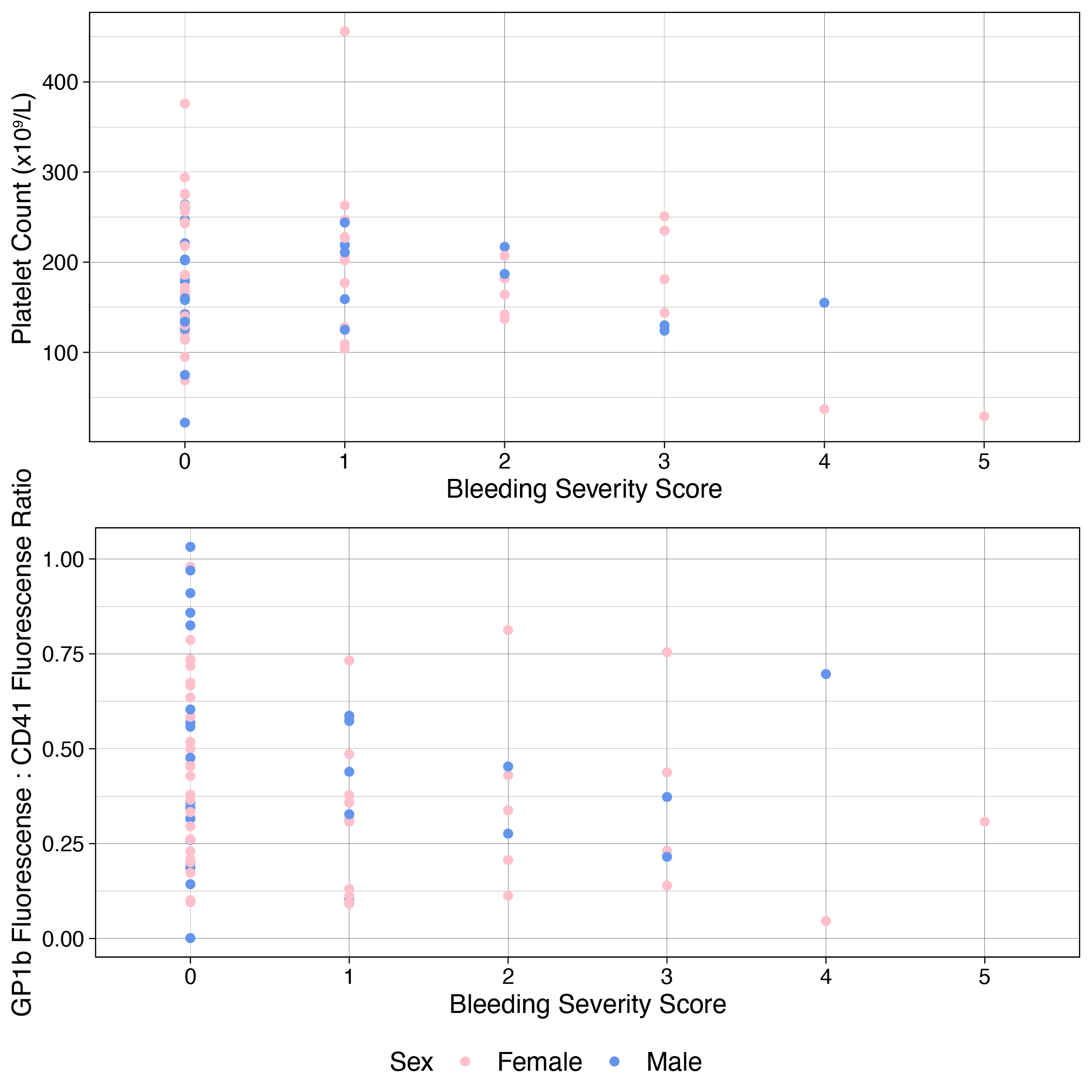

**Supplementary Figure 5:** Platelet parameters versus bleeding score. (A) comparison of bleeding score and platelet count. (B) comparison of GP1b : CD41 ratio versus bleeding score. Pink points are female participants while blue represents males.
